# Classification of Coronavirus Images using Shrunken Features

**DOI:** 10.1101/2020.04.03.20048868

**Authors:** Saban Ozturk, Umut Ozkaya, Mucahid Barstugan

## Abstract

Necessary screenings must be performed to control the spread of the Corona Virus (COVID-19) in daily life and to make a preliminary diagnosis of suspicious cases. The long duration of pathological laboratory tests and the wrong test results led the researchers to focus on different fields. Fast and accurate diagnoses are essential for effective interventions with COVID-19. The information obtained by using X-ray and Computed Tomography (CT) images is vital in making clinical diagnoses. Therefore it was aimed to develop a machine learning method for the detection of viral epidemics by analyzing X-ray images. In this study, images belonging to 6 situations, including coronavirus images, are classified. Since the number of images in the dataset is deficient and unbalanced, it is more convenient to analyze these images with hand-crafted feature extraction methods. For this purpose, firstly, all the images in the dataset are extracted with the help of four feature extraction algorithms. These extracted features are combined in raw form. The unbalanced data problem is eliminated by producing feature vectors with the SMOTE algorithm. Finally, the feature vector is reduced in size by using a stacked auto-encoder and principal component analysis to remove interconnected features in the feature vector. According to the obtained results, it is seen that the proposed method has leveraging performance, especially in order to make the diagnosis of COVID-19 in a short time and effectively.

## 1. INTRODUCTION

The Coronavirus (COVID-19), which appeared towards the end of 2019, caused a global epidemic problem that could spread quickly from the individual to the individual in the community. According to the World Health Organization (WHO) data, the rate of catching COVID-19 in China is between 16-21%, and the mortality rate is 2-3% [1]. For this reason, people need to take the necessary quarantine measures and implement protection procedures to deal with COVID-19. The clinical symptom must be present, and also positive X-ray and CT images must be included with the positive pathological test to be able to diagnose COVID-19. Fever, cough, and shortness of breath are frequently seen as clinical symptoms [2]. Besides, positive findings should be obtained in X-ray and CT images with these symptoms. Another diagnostic method of COVID-19 is to examine the RNA sequence of the virus. However, this method is not a very efficient technique due to a long time of diagnosis. Also, its accurate diagnosis rate varies between 30-50%. Thus, diagnosis tests should be repeated in lots of time [3]. Radiological imaging techniques are essential for the detection of COVID-19. In X-rays and Computer Tomography (CT) images, the COVID-19 virus shows the same features in the early and late stages. Although it shows a circular distribution within an image, it may exhibit similar characteristics with other viral epidemic lung diseases [4]. This makes it challenging to detect COVID-19 from other viral cases of pneumonia.

Machine learning techniques, which are a sub-branch of the field of artificial intelligence, are frequently used for medical applications in the concept of feature extraction and image analysis. Machine learning methods are used in the diagnosis of viral pneumonia, especially in X-ray and CT images, tumor diagnosis, and cystoscopic image analysis [5]. Viral pathogenic patterns contain several features in X-ray and CT images [6]. COVID-19 shows an irregular distribution and shading within the image [7]. When the X-ray and CT studies in the literature are examined, it is seen that hand-crafted, and recently CNN-based methods are used [8, 9]. Studies with hand-crafted features are generally seen in studies performed before the CNN method. Studies with hand-crafted features are generally seen in studies performed before the CNN method. These studies are highly dependent on the results produced by the methods chosen based on the researcher’s experience. In addition, it is not always possible to achieve the same performance for different datasets. These methods cover many techniques from edge detection algorithms to GLRLM and SFTA methods. Sorensen et al. [10] used dissimilarities computed between collections of regions of interest. Then, they classified these features via a standard vector space-based classifier. Zhang and Wang [11] presented a CT classification framework with three classical types of features (grayscale values, shape and texture features, and symmetric features). They used the radial basis function of the nerve network to classify image features. Homem et al. [12] presented a comparative study using the Jeffries–Matusita (J–M) distance and the Karhunen–Loève transformation feature extraction methods. Albrecht et al. [13] proposed a classification framework with an average greyscale value of images for multi-class image classification. Yang et al. [14] suggested an automatic classification method to classify breast CT images using morphological features. When these studies with hand-crafted features and other similar studies in the literature are examined, it is seen that the proposed methods are mostly successful only on one dataset. Performance decreases when the same operation is done with other datasets.

In addition, interest in hand-crafted methods has begun to decline with the introduction of CNN and other automated feature extraction techniques. Convolutional neural network architecture is a deep learning architecture that automatically extracts and classifies images from images [15]. Özyurt et al. [16] proposed a hybrid model called fused perceptual hash-based CNN to reduce the classifying time of liver CT images and maintain performance. Xu et al. [17] used a transfer learning strategy to deal with the medical image unbalance problem. Then, they compared GoogleNet, ResNet101, Xception, and MobileNetv2 performances. Lakshmanaprabu et al. [18] analyzed the CT scan of lung images using the assistance of optimal deep neural network and linear discriminate analysis. Gao et al. [19] converted raw CT images to low attenuation, raw images, and high attenuation pattern rescale. Then, they resampled these three samples and classified them using CNN.

The methods in the literature have various drawbacks. When CT studies with hand-crafted features are examined, it is seen that the one-way features in these studies show high performance only in the dataset of interest. It also creates a computational load. On the other hand, in CNN studies, a rather sizeable labeled dataset is needed. Unbalanced and unlabeled data pose problems for CNN education. Besides, it requires a high hardware capacity.

In this study, epidemic classification is made from X-ray and CT images by extracted hand-craft features. Images in the dataset used to consist of ARds, Covid, No finding, pneumocystis-pneumonia, Sars, and streptococcus classes. As it is known, since Covid is a very new disease, the data obtained are quite limited. In this case, the dataset is not suitable for using CNN. Also, the dataset is unbalanced. Therefore, this will cause poor performance not only when using the dataset with CNN, but also with hand-crafted features. Although similar problems are solved in the literature with the transfer learning approach, transfer learning studies are not suitable for the medical domain. To solve all these problems, feature vectors are created in different spatial planes using four hand-crafted feature extraction algorithms. These four feature vectors created for each image are combined into a single vector. To solve the unbalanced dataset problem, image augmentation and data over-sampling are performed to reproduce the missing number of class vectors. The reason for using these two methods is to prevent the synthetic performance effect of synthetic data generated by the SMOTE algorithm. When classified with extended features for a small number of observations, noises and various irrelevant information act as disruptors.

For this reason, the size of the feature vector is reduced by using a stacked auto-encoder (sAE) and principal component analysis (PCA). The success of these two feature reduction methods, which work according to different approaches, is compared. Thus, both storage space is saved, and response time is accelerated. Finally, data is classified with the support vector machine (SVM).

This paper is organized as follows. Section 2 describes details about the dataset, the introduction of the proposed framework method, and parameters. Section 3 presents experiments and experimental results. Section 4 includes discussion, and Section 5 presents the conclusion.

## 2. MATERIAL and METHODS

### 2.1. Dataset Description

In the chest X-ray and CT images of patients with COVID-19, there are medium-characteristic infected patterns [20]. Accurate analysis of positive and negative infected patients is essential to minimize the rate of spread of COVID-19. At the same time, the pre-recognition system should have a low level of false-positive alarms to serve more patients. Images in the dataset^1^ used to consist of ARds, Covid, No finding, pneumocystis-pneumonia, Sars, and streptococcus classes. Dataset includes: 4 ARds images, 101 Covid images, 2 No finding images, 2 pneumocystis-pneumonia images, 11 Sars images, and 6 streptococcus. Figure 1 contains some sample images from the dataset.

**Figure 1.**
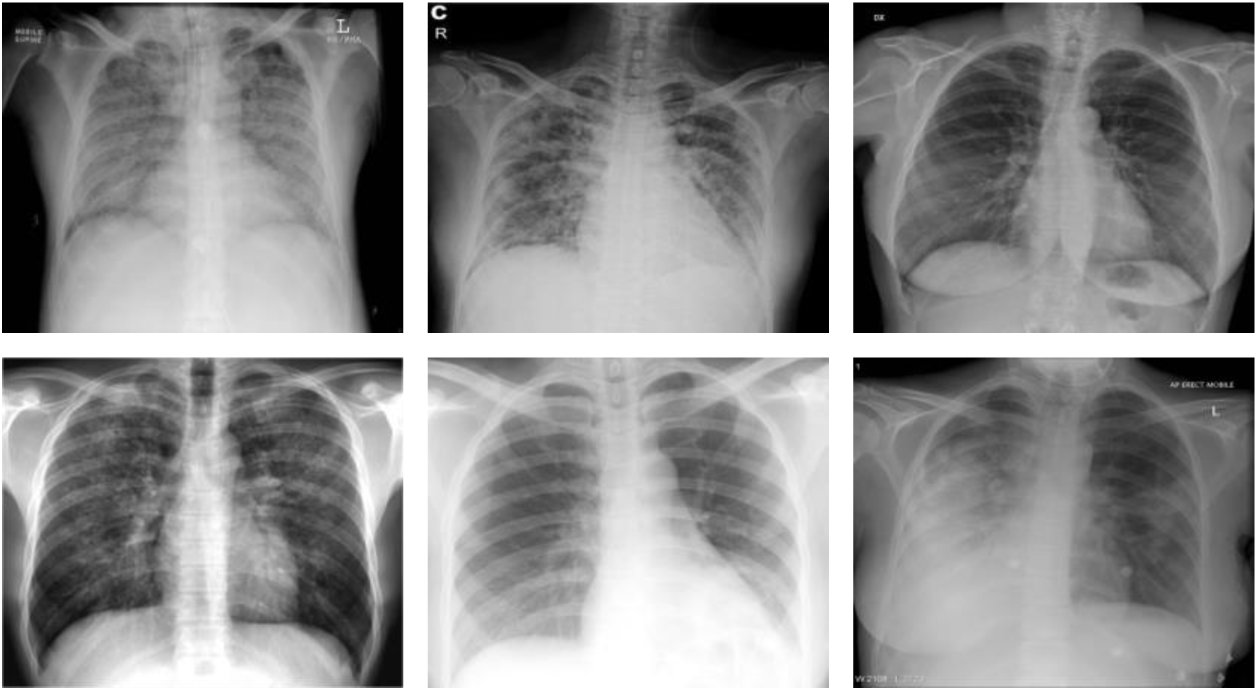
Sample images from the dataset.

### 2.2. Proposed Method Overview

The proposed framework is shown in Figure 2. Firstly, classical image augmentation is applied to minority classes, and images are rotated, scaled, etc. The total number of images from after image augmentation 126 increases to 260.

**Figure 2.**
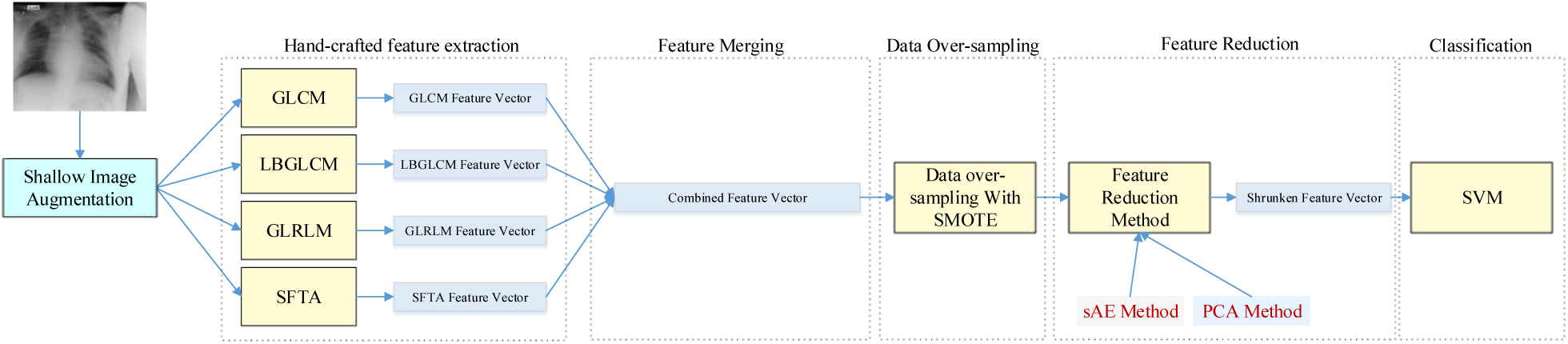
Proposed framework overview.

Then, four hand-crafted features are extracted from all images. By combining these feature vectors, 78 features are obtained for each image. Then the feature vectors of 260 images consisting of 78 features are over-sampling with the SMOTE method. After this process, 495 feature vectors are created. With these feature vectors, sAE and PCA are trained, respectively. The purpose of the sAE and PCA algorithms in this study are to narrow down 78 features and obtain 20 features. Finally, SVM is trained with 495 vectors containing 20 features for classification purposes. The necessity of both image augmentation and data over-sampling arises from the depth of the unbalanced structure in the dataset. In case only image augmentation is applied, there are only two images in many classes, and almost the same images will be produced. In this case, in-class overfitting occurs. When using only synthetic data over-sampling method, synthetic performance data is obtained, and performance may remain low in real applications. For this reason, two data replication techniques are combined.

#### 2.2.1. The Feature Extraction Techniques

Grey Level Co-occurrence Matrix (GLCM), Local Binary Grey Level Co-occurrence Matrix (LBGLCM), Grey Level Run Length Matrix (GLRLM), and Segmentation-based Fractal Texture Analysis (SFTA) features have been extracted to classify pandemic diseases.

*Grey Level Co-occurrence Matrix;* many statistical features from a grey level image form GLCM as shown in Figure 3. Features square matrix is defined as *G(i,j)*. Four different directions are used to divide the *G* matrix into normalized typical formation matrices. These directions are defined as vertical, horizontal, left, and right cross directions. These are computed for each of these adjacent directions. These texture features are defined as angular secondary moment, contrast, correlation, the sum of squares, variance, inverse difference moment, sum average, sum variance, sum entropy, entropy, difference entropy, difference variance, information measures of correlation 1, information measures of correlation 2, autocorrelation, dissimilarity, cluster shade, cluster prominence, maximum probability, and the inverse difference [21].

**Figure 3.**
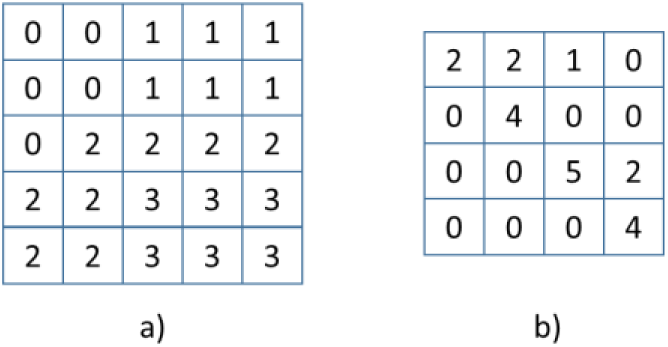
a) Grey Level Images b) GLCM matrix for direction 0^0^ with 1 distance.

*Local Binary Grey Level Co-occurrence Matrix; the LBGLCM feature extraction method is a hybrid* technique used together with Local Binary Pattern (LBP) and GLCM. LBP technique is applied to the grey-level image. Then, GLCM features are extracted from the obtained LBP texture image. GLCM method takes into account neighboring pixels at the feature extraction stage. It does not perform any operation on other local patterns in the image. Textural and spatial information in the image is obtained together with the LBGLCM method. Simultaneous acquisition of this information increases the availability of the LBGLCM algorithm in image processing applications [22].

*Grey Level Run Length Matrix;* GLRLM uses higher-order statistical methods to extract the spatial features of gray level pixels. The obtained feature matrix is two-dimensional. Each value in the matrix shows the total formation value of the gray level. GLRLM features are seven in total. These high statistical features are the short-run emphasis, long-run emphasis, gray-level non-uniformity, run-length non-uniformity, run percentage, low gray-level run emphasis, and high gray-level run emphasis [23].

*Segmentation-based Fractal Texture Analysis;* In texture analysis, low computing time and efficient feature extraction are critical. SFTA technique is a method that can be evaluated in this concept. In the SFTA method, the image is converted into a binary structure by multiple thresholding technique. Thresholding values of *t*_*1*_, *t*_*2*_, *t*_*3*_,.. *t*_*n*_ are performed. Interclass and in-class variance values are used to determine the threshold sets. In order to minimize the in-class variance value, the optimum threshold number is applied to the image regions.

Figure 4 shows the feature extraction stages of the Pseudocode SFTA algorithm. *V*_*SFTA*_ represents the obtained feature vector. Initially, multiple threshold values (*T*), all pairs of contiguous thresholds (*T*_*A*_), and threshold values (*T*_*B*_) corresponding to maximum grey levels are determined. Then, segmented images pixels, borders, and *V*_*SFTA*_ are updated for all threshold values in a loop. The asymptotic complexity of the obtained *V*_*SFTA*_ vector is *O(N·*| *T*| ). While *N* expresses the number of pixels, | *T*| shows the number of different thresholds resulting from the multi-level Otsu algorithm [24].

**Figure 4.**
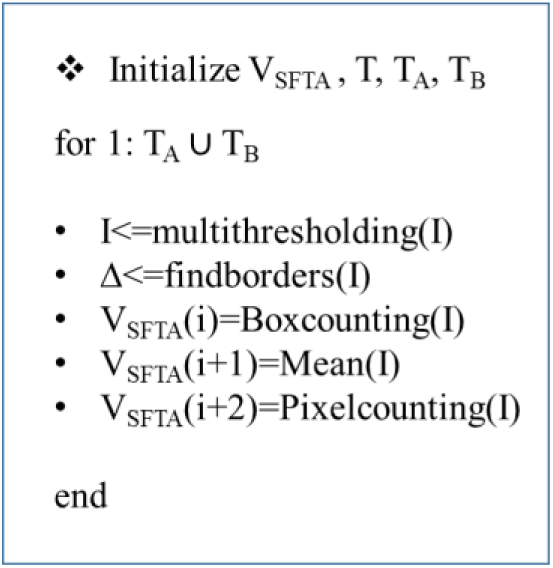
Pseudo Code of SFTA.

#### 2.2.2. Over-sampling with SMOTE

Synthetic Minority Over-sampling Technique (SMOTE) is useful for overcoming unbalanced class distribution problems for classification tasks [25]. The SMOTE algorithm increases the number of samples related to the minority class by producing synthetic samples. To over-samples minority class data is used at a certain rate. This ratio can be selected differently for each class. Let *X=[X*_*1*_, *X*_*2*_, *…, X*_*n*_] be the feature vectors of each class. The total number of classes is represented by *n*. If the minority class is represented by *X*_*minor*_, synthetic points are created by determining data points about *X*_*minor*_. *K* neighborhood value is used to determine data points. According to K-nearest neighbors, Equation 1 is used to calculate a new sample.

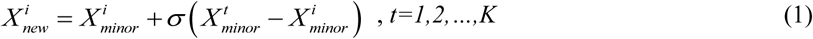

where 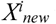 represents the new synthetic sample. *σ* represents a random number uniformly distributed within the range [0,1]. In the use of this study, the highest number of samples is determined to create an almost equal number of samples for all classes. It is calculated according to the number of samples in other classes. As the upper limit, eight times the number of samples in the minority class is selected. This criterion applies only to examples in the no finding class.

#### 2.2.3. Stacked Auto-encoder

An AE architecture consists of two layers, encoder, and decoder, whose main purpose is to re-interpret the relationship between input and output. In the proposed study, AE architecture expresses the high-dimensional feature vector with fewer parameters. AE architecture, trained in an unsupervised style, creates a relationship between input and output. In AE architecture, more than one AE is added in series. Let *X={x*_*n*_*}*^*N*^ be an N feature vector. New shrunken feature vectors to be obtained at the sAE output *F:X→[0,1]*^*Nxk*^, is a k-bit vector (*b*_*n*_*ϵ[0,1]*^*k*^) for each feature vector *x*_*n*_. The output of the AE layer is calculated as in Equation 2.

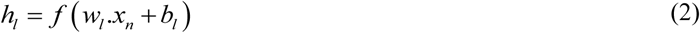

where *h*_*l*_ is the output, *w*_*l*_ represents weights, *x*_*n*_ is *n*th feature vector, *f* is an activation function, and *b*_*l*_ represents bias parameter. *f(x)=1/(1+e*^*-x*^) is used as activation function. The proposed sAE method produces feature vectors consisting of 50 feature features and 50 feature features. The sAE part of our framework is shown in Figure 5.

**Figure 5.**
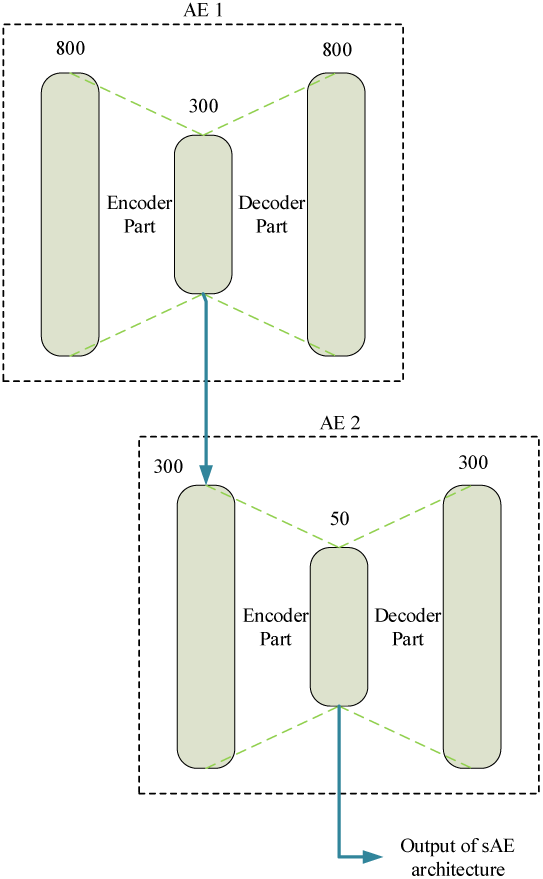
The sAE part of the proposed framework.

#### 2.2.4. Principle Component Analysis (PCA)

The main purpose of Principle Component Analysis (PCA) is to extract the most significant features from the available data. In this way, it ensures that many feature variables are reduced without any loss of information. PCA takes its place in the literature as a linear analysis method. A different coordinate system occurs by rotating the linear combinations of p randomly distributed data (x1, x2,…, xp) around the original axis. The axes in the new coordinate system show the directions of the highest variability. The primary purpose of performing this coordinate conversion is to provide a better interpretation of the data. In cases where the correlations are quite evident in feature reduction, different spinning techniques may show similar results [26]. Obtained features after the rotation are more meaningful.

#### 2.2.5. Support Vector Machines (SVMs)

Support Vector Machines (SVMs) can classify the data with the help of planes. The borders between classes are determined according to these planes. The plane between objects creates a boundary between classes. Linear planes may not show high performance in the classification process. Therefore, it can be necessary to use nonlinear parabolic hyperplanes. Parabolic hyperplanes can be capable of inter-class problem. The mentality of the SVMs algorithm is visualized in Figure 6. Features are transferred to a different space by using kernel functions. This process is defined as conversion or matching. Thus, features can be distinguished by linear planes in the new space [27].

**Figure 6.**
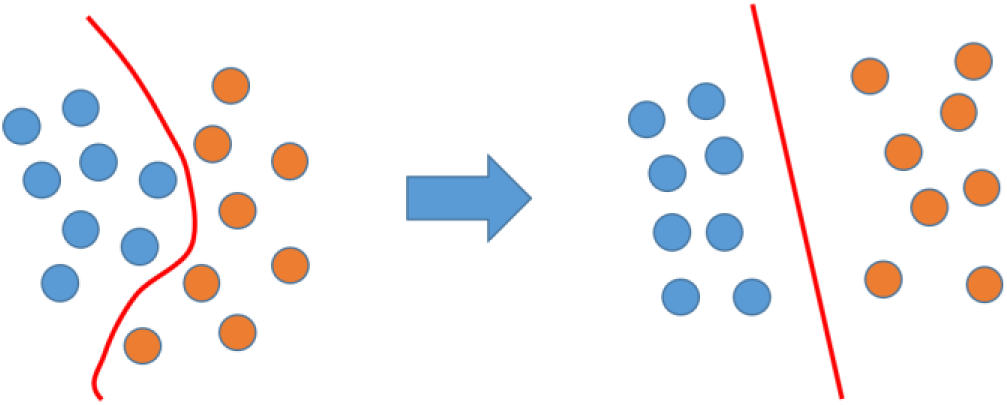
Conversion or Matching Process.

## 3. EXPERIMENTAL RESULTS

The proposed method is trained on a computer with Intel Core i7-7700K CPU (4.2 GHz), 32 GB DDR4 RAM, and NVIDIA GeForce GTX 1080 graphic card.

The dataset used consists of six classes: ARds, Covid, No finding, pneumocystis-pneumonia, Sars, and streptococcus classes. It contains 126 images in total, including 4 ARds images, 101 Covid images, 2 No finding images, 2 pneumocystis-pneumonia images, 11 Sars images, and 6 streptococci. When the image numbers are examined, it is understood that the dataset is a unique dataset for the detection of the Covid virus. However, the numbers of other classes in this dataset cause an overfitting problem in the classification process. For this reason, the number of minority classes should be increased. Because clinical studies related to Covid have just started and the difficulty of labeled data access, it seems impossible to obtain sufficient Covid data for CNN training.

For this reason, the feature extraction method is most suitable for hand-crafted methods. However, the fact that the number of images in the dataset is quite low and the number of sample differences between classes is very high (almost 90% belong to only one class) will create a problem for classifier algorithms. For this purpose, the number of images and data are increased with the image augmentation technique and data over-sampling. Firstly, the number of images in the dataset is increased at the highest possible level, and 260 images in total are obtained. After image augmentation, it consists of a total of 260 images, including 40 ARds, 101 Covid, 24 No findings, 24 pneumocystis-pneumonia images, 43 Sars, 28 streptococcus images.

Hand-crafted features are extracted from 260 images created using GLCM, LBGLCM, GLRLM, and SFTA feature extraction algorithms. In this case, by selecting the GLCM algorithm offset parameter [2 0], 22 features are extracted for each image, by selecting the LBGLCM algorithm offset parameter [2 0], 22 features are extracted, the GLRLM algorithm generates 7 features, and when the SFTA algorithm feature parameter is selected as 5, it generates 27 features (*6*feature_parameter-3*). When feature vectors of each image are combined, feature vectors consisting of 78 features of each image is obtained. Feature vectors of 260 images consisting of 78 features are increased by using the SMOTE algorithm. According to 6 neighborhood values, the SMOTE algorithm is used. Accordingly, the numbers of over-sampled class samples are as follows; for the ARds class, the multiplication coefficient is selected 2 and 80 samples are obtained, for the Covid class, the multiplication coefficient is selected 0 and 101 samples are obtained, for the No findings class, the multiplication coefficient is selected 3 and 72 samples are obtained, for the pneumocystis-pneumonia class, the multiplication coefficient is selected 3 and 72 samples are obtained, for the sars class, the multiplication coefficient is selected 2 and 86 samples are obtained, for the streptococcus, the multiplication coefficient is selected 3 and 84 samples are obtained. In this case, the total number of samples is 495.

### 3.1. Classification Results of Raw Feature Vectors

The proposed framework includes three stages: feature extraction, over-sampling, and shrunken features. For this purpose, experiments are carried out in these three stages. Firstly, the classification results with 260 samples and 495 samples are examined for 78 features in raw form. The contribution of the over-sampling method is investigated by comparing the performance of the classification processes with the SVM algorithm. Table 1 shows the performance parameters of raw feature vectors consisting of 78 features of 260 samples and raw feature vectors of 495 samples created with the SMOTE algorithm. It is seen that increasing the minority classes with synthetic samples has a positive effect on classification accuracy.

**Table 1.** The classification results of raw feature vectors

| Methods | Sensitivity | Specificity | Accuracy | Precision | F-Score |
| --- | --- | --- | --- | --- | --- |
| Raw features with 260 samples | 75.4% | 94.94% | 76.92% | 74.07% | 73.25% |
| Raw features with 495 samples | 83.15% | 96.96% | 86.54% | 86.35% | 84.55% |

In Table 1, it is seen that the classification made with the addition of synthetic classes creates a performance contribution of more than 10%. One of the most important reasons for this is the elimination of imbalance between classes. Another reason is that the number of samples almost doubles. The AUC curves of the experiments are shown in Figure 7.

**Figure 7.**
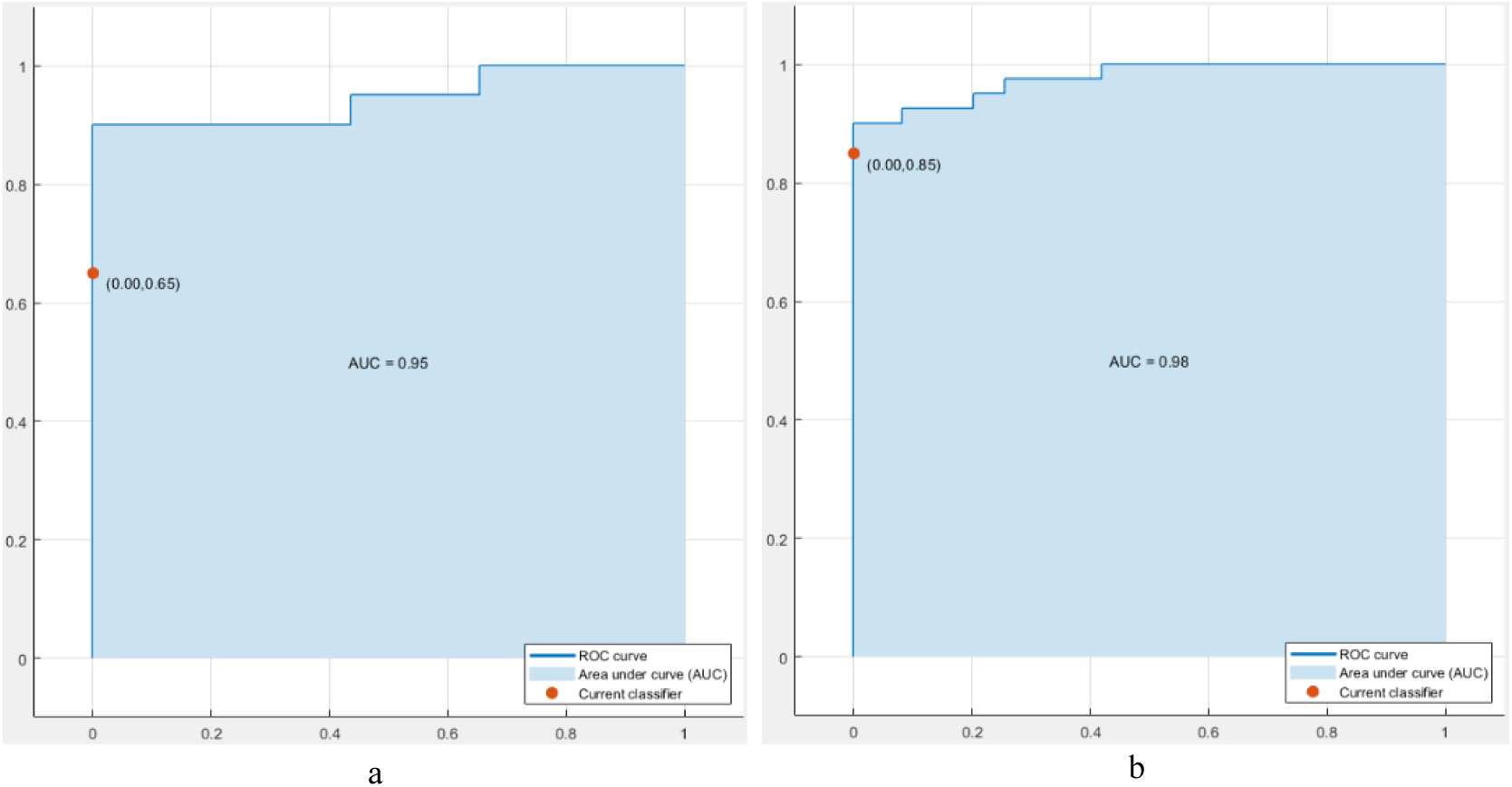
AUC curves of raw features, a) 260 samples, b) 495 samples.

### 3.2. Classification Results of sAE

The feature vector in the raw state is quite long, and long feature vectors pose a variety of problems for a limited number of observations. The most important of these is the learning of noises and the problem of sticking to irrelevant points of interest. Other problems are the storage problem and computational complexity. To overcome these problems, the feature vector is narrowed by the sAE method. In this method, which we call shrunken features, sAE reduces the length of the high-length feature vectors. 20 features are obtained at the recommended sAE output. With these features, two experiments were carried out. In the first experiment, only the results related to 260 samples reproduced with image augmentation, and in the second experiment, 495 sample results obtained with the SMOTE algorithm are examined. Table 2 shows the classification performances obtained using SVM.

**Table 2.** The classification results of shrunken features with sAE

| Methods | Sensitivity | Specificity | Accuracy | Precision | F-Score |
| --- | --- | --- | --- | --- | --- |
| shrunk features with 260 samples | 61.04% | 92.65% | 65.77% | 62.62% | 61.45% |
| shrunk features with 495 samples | 68.91% | 93.91% | 71.92% | 69.89% | 69.13% |

When Table 1 and Table 2 are compared, it is seen that the classification performance decreases considerably. It is understood that education leads to memorization in sAE architecture with its low number of samples and synthetic data. As seen in the AUC curves in Figure 8, the sAE method cannot be used with these data due to the overfitting problem.

**Figure 8.**
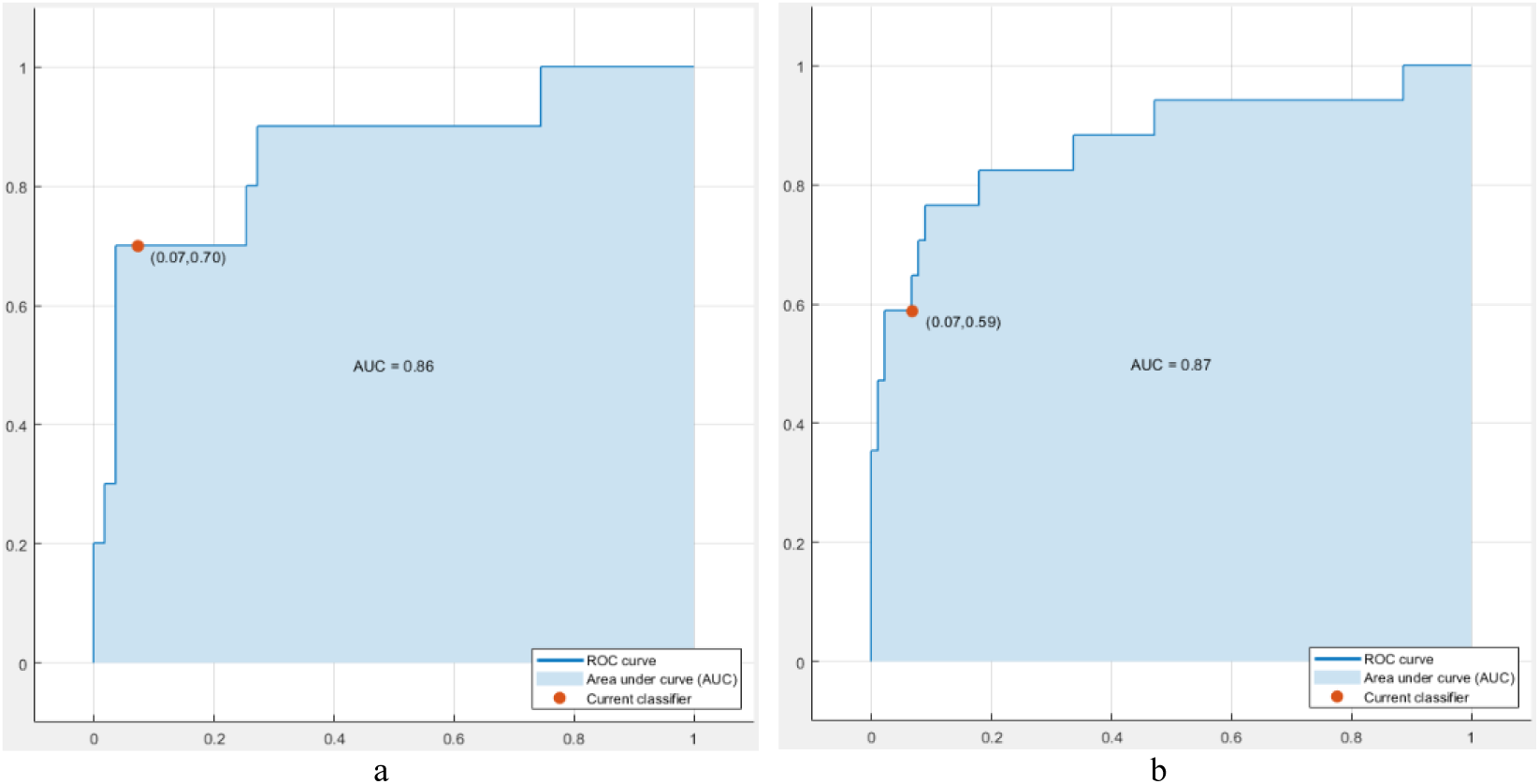
AUC curves of shrunken features with sAE, a) 260 samples, b) 495 samples.

### 3.3. Classification Results of PCA

Since the sAE method is supervised, it is understood that feature narrowing operation with the insufficient number of samples has failed. For this reason, the PCA algorithm, which performs feature reduction in unsupervised style, is tried. PCA algorithm transforms 78 features to 20 features according to their relations. To examine the balanced dataset effect, classification experiments are applied to the PCA algorithm with 260 samples and 495 samples. Classification accuracy and AUC results obtained are shown in Table 3. Comparing these results with Table 1 and Table 2, it is understood that the performance of the PCA algorithm is higher. Considering the proposed framework and dataset status, it is seen that the PCA algorithm is more suitable for feature selection. It is thought that its effect will be high, especially in the investigation of viruses such as Covid that started suddenly and in studies with a small number of data.

**Table 3.** The classification results of shrunken features with PCA

| Methods | Sensitivity | Specificity | Accuracy | Precision | F-Score |
| --- | --- | --- | --- | --- | --- |
| shrunken features with 260 samples | 85.08% | 97.32% | 88.46% | 89.75% | 87.12% |
| shrunken features with 495 samples | 91.88% | 98.54% | 94.23% | 96.73% | 93.99% |

In Table 3, it is seen that in experiments with 260 samples, it produces more successful results than other experiments, regardless of the number of features. After over-sampling, the classification performance increases even more due to the class balance. Figure 9 shows the AUC curve of the features obtained by the PCA method.

**Figure 9.**
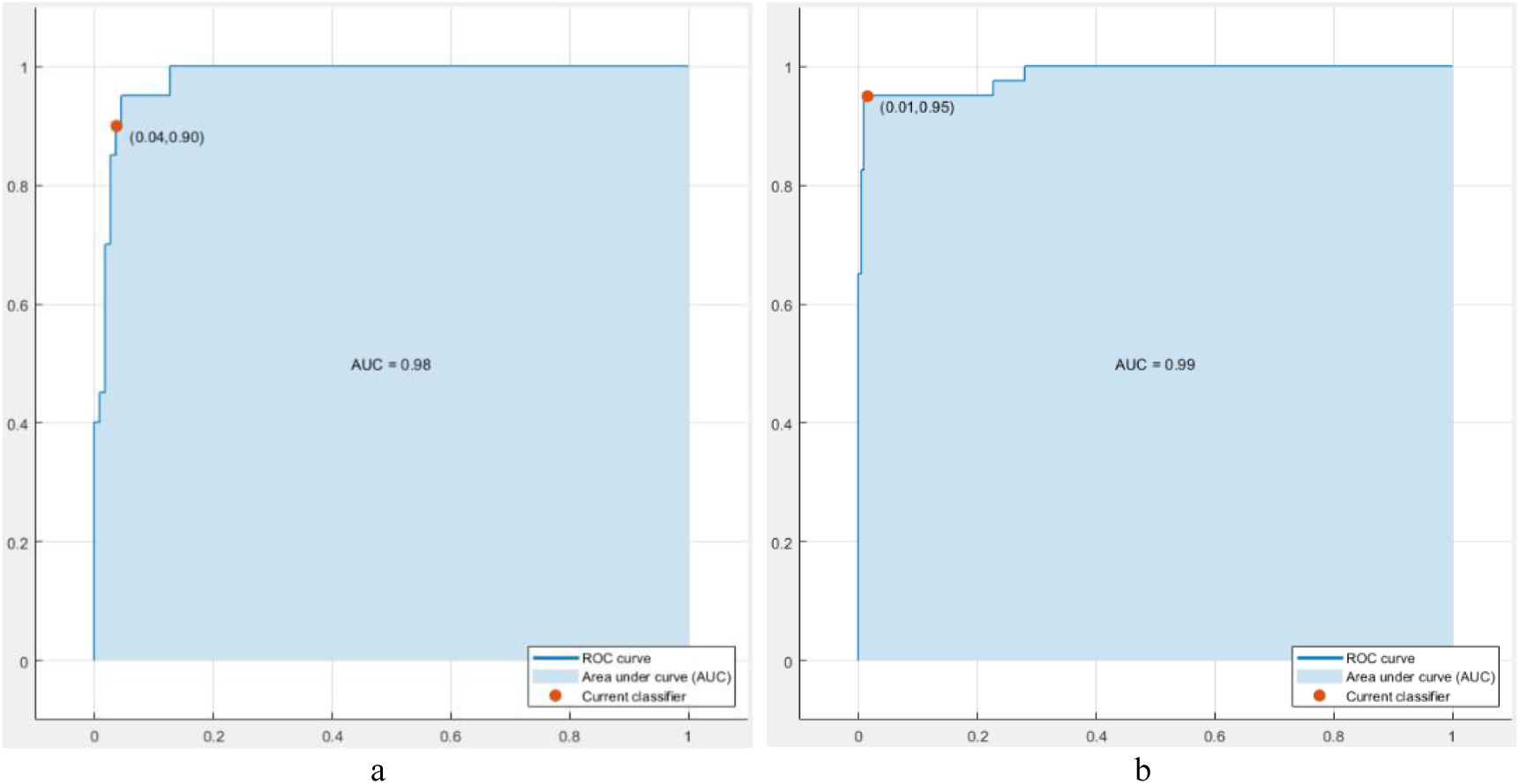
AUC curves of shrunken features with PCA, a) 260 samples, b) 495 samples.

## 4. DISCUSSION

The proposed method aims to detect sudden outbreaks such as Coronavirus automatically. However, in such cases, it is difficult to find a sufficient number of labeled data. In this case, it is shown that 90% of accuracy performances can be achieved by using hand-crafted features rather than using CNN-based methods. In addition, the contributions of image augmentation and data over-sampling methods are examined for datasets, where the number of samples between classes is quite unstable. As can be seen from the second lines of Table 1, Table 2, and Table 3, the contribution of these methods to performance is almost 10%. Of course, the similarities between real data and synthetic data are quite high, but synthetic data generally only uses the in-class variation.

For this reason, classifier performance can be misleading. However, when all comparisons are examined, the common contribution cannot be ignored. The results of the sAE method, which produces favorable results for many studies in the literature, are surprising. The main reasons for this are the low number of samples, the imbalance between classes, and the closeness in synthetic data. When this situation causes in-class affinity, it creates a code generation problem for sAE. It appears that it is not appropriate to use a CNN architecture for the training of such datasets, which is insufficient even for a shallow sAE architecture. Similar to the approach to hand-crafted feature extraction techniques, PCA architecture was considered instead of the sAE architecture. Since the PCA architecture can operate independently from the number of samples, it has provided very successful classification performance.

As a result, it is not often appropriate to use today’s methods in the supervised style to examine newly emerged datasets containing an insufficient number of samples. Similarly, datasets with an unbalanced class structure are not suitable for training. In cases where it is not possible to find or wait for more labeled data, these datasets can be successfully represented by hand-crafted features. For the unbalanced class problem, performing two-stage data replication instead of a single-sided data replication provides higher performance.

## 5. CONCLUSION

Early diagnosis and control of infectious diseases such as COVID-19 are vital for public health. Therefore, automated pre-diagnosis systems are needed to help diagnose the disease quickly and accurately. In this study, we developed a machine learning-based system that can analyze both X-ray and CT images. These days when the access to the Corona data is very limited, experiments are carried out with a dataset with very little data and inter-class imbalance. Due to the limitations of the dataset, hand-crafted methods are preferred over deep learning-based methods. The proposed framework performance, which is based on combining feature vectors produced by four hand-crafted methods and then reproducing with over-sampling and augmentation methods, is very inspiring. It produces beneficial results, especially in terms of comparing sAE and PCA performances. The results of the sAE and PCA algorithms are presented simultaneously, to provide the study to be useful for the researchers who want to work in this field. Besides, the image augmentation and data over-sampling effect are demonstrated in experiments. In future studies, a broader dataset will be produced with more balanced and more labeled Covid-19 data. Also, CNN architectures that can produce a high performance on such datasets will be tried to be developed.

## Data Availability

The dataset is shared as a supplemental file.

## Compliance with Ethical Standards

### Human and animal rights

The paper does not contain any studies with human participants or animals performed by any of the authors.

## Footnotes

1 https://github.com/ieee8023/covid-chestxray-dataset

